# COVID-19 induces a hyperactive phenotype in circulating platelets

**DOI:** 10.1101/2020.07.24.20156240

**Authors:** Shane P. Comer, Sarah Cullivan, Paulina B. Szklanna, Luisa Weiss, Steven Cullen, Sarah Kelliher, Albert Smolenski, Niamh Moran, Claire Murphy, Haidar Altaie, John Curran, Katherine O’Reilly, Aoife G. Cotter, Brian Marsh, Sean Gaine, Patrick Mallon, Brian McCullagh, Fionnuala Ní Áinle, Barry Kevane, Patricia B. Maguire, On behalf of the COCOON Study investigators

**Author notes:** **Corresponding authors**: Prof Patricia B. Maguire, Conway SPHERE Research Group, UCD Conway Institute, University College Dublin, Belfield, Dublin 4, Ireland, Dr Barry Kevane, Conway SPHERE Research Group, Department of Haematology, Mater Misericordiae University Hospital, Eccles Street, Dublin 7, Ireland., Prof Fionnuala Ní Áinle, Conway SPHERE Research Group, Department of Haematology, Mater Misericordiae University Hospital, Eccles Street, Dublin 7, Ireland. These authors contributed equally to this manuscript (Shane P. Comer, Sarah Cullivan, Paulina B. Szklanna, Luisa Weiss).

## Abstract

**Background:** Coronavirus disease 2019 (COVID-19), caused by novel coronavirus SARS-CoV-2, has to date affected over 13.3 million globally. Although high rates of venous thromboembolism and evidence of COVID-19-induced endothelial dysfunction have been reported, the precise aetiology of the increased thrombotic risk associated with COVID-19 infection remains to be fully elucidated.

**Objectives:** Here, we assessed clinical platelet parameters and circulating platelet activity in patients with severe and non-severe COVID-19.

**Methods:** An assessment of clinical blood parameters in patients with severe COVID-19 disease (requiring intensive care), patients with non-severe disease (not requiring intensive care), general medical in-patients without COVID-19 and healthy donors was undertaken. Platelet function and activity were also assessed by secretion and specific marker analysis.

**Results:** We show that routine clinical blood parameters including increased MPV and decreased platelet:neutrophil ratio are associated with disease severity in COVID-19 upon hospitalisation and intensive care unit admission. Strikingly, agonist-induced ADP release was dramatically higher in COVID-19 patients compared with non-COVID-19 hospitalized patients and circulating levels of PF4, sP-selectin and TPO were also significantly elevated in COVID-19.

**Conclusion:** Distinct differences exist in routine full blood count and other clinical laboratory parameters between patients with severe and non-severe COVID-19. Moreover, we have determined that COVID-19 patients possess hyperactive circulating platelets. These data suggest that abnormal platelet reactivity may contribute to hypercoagulability in COVID-19. Further investigation of platelet function in COVID-19 may provide additional insights into the aetiology of thrombotic risk in this disease and may contribute to the optimisation of thrombosis prevention and treatment strategies.

**Essentials:**

- Routine platelet-related clinical blood parameters (MPV, PNR) are associated with disease severity in COVID-19.
- Agonist-induced ADP release is dramatically higher in COVID-19 patients compared with non-COVID-19 hospitalized patients.
- Circulating levels of PF4, sP-selectin levels and TPO are significantly elevated in COVID-19.
- Identification of a hyperactive platelet phenotype may warrant re-evaluation of current thrombotic prevention strategies in COVID-19 treatment.

## Introduction

Over 13.3 million people globally have been infected since the outbreak of severe acute respiratory syndrome-coronavirus-2 (SARS-CoV-2; or COVID-19) in December 2019, with over 578,000 fatalities reported to date (1). COVID-19 is characterised by a marked pro-inflammatory response with fever, elevated inflammatory markers and clinical & radiological features of pneumonitis being evident among affected individuals (2). A complex interplay is known to exist between pro-inflammatory pathway activity and blood coagulation activation; this interplay appears to represent a source of morbidity among SARS-CoV-2 infected patients, particularly among those with severe disease (3). Unexpectedly high levels of venous thromboembolism (VTE) (4-10) have been reported in this patient group and levels of D-dimer (a marker of increased blood coagulation activation) have been shown to correlate with disease severity and appear to be predictive of mortality (11, 12). COVID-19 patients with cardiovascular risk factors (hypertension, diabetes) have been indentified as being particularly at risk of thrombotic events (13-16). Moreover, recent post-mortem studies have shown evidence of widespread thrombosis in pulmonary vasculature and other organs (17, 18).

Anti-platelet drugs (such as aspirin) and anti-coagulants (such as low molecular weight heparin (LMWH)) are known to be effective in the prevention and treatment of arterial and venous thrombosis (19-21). High rates of VTE despite LMWH thromboprophylaxis have been reported in COVID-19 and emerging reports of unusual or severe arterial thrombotic events have been published (5, 18, 22-24). As both arterial and venous thrombosis can have devastating consequences, efforts to determine the precise molecular mechanisms underlying the unique hypercoagulable state in COVID-19 must be prioritised in order to optimise preventative and therapeutic strategies (25, 26).

Circulating platelets, the activity of which are central to haemostasis and thrombosis, are now understood to fulfill a much broader role in the regulation of a myriad of biological processes including inflammation, wound healing and angiogenesis (27). Platelet-mediated immune responses induced by viral infection have previously been reported (28-31) and a recent report has demonstrated altered platelet gene expression profiles and functional responses in patients infected with COVID-19 (32). Moreover, thrombocytopenia has been noted in response to dengue (33, 34) and SARS-CoV infection (35) and among in-patients with COVID-19, thrombocytopenia appears to be associated with increased risk of in-hospital mortality (3, 36). Increased mean platelet volume (MPV), a sensitive indicator of circulating platelet activity and a prognostic marker in thromboinflammation (37-39), has also been reported in association with specific viral infections (40).

In the present study, we aimed to characterise patterns of platelet activity among patients infected with COVID-19 relative to that observed in non-affected hospitalised patients in order to determine if platelet dysfunction or hyperactivity contributes to COVID-19-induced hypercoagulability.

## Methods

### Patient recruitment and blood collection

Ethical approval to proceed with this study (1/378/2077) was granted by the Institutional Review Board of the Mater Misericordiae University Hospital. Anonymised datasets describing clinical laboratory parameters among hospitalised patients with severe COVID-19 (requiring critical care support), non-severe COVID-19 (not requiring critical care) and non-COVID-19 affected medical inpatients were compiled from routine clinical testing results. SARS-CoV-2 infection was confirmed in all cases by RT-PCR analysis of nasopharyngeal swab specimens. Informed consent was sought prior to obtaining additional blood samples for analysis from a sub-group of patients with severe COVID-19, non-severe COVID-19 and hospitalised patients without COVID-19, with patients matched for age, body mass index (BMI) & gender. All patients were recruited at the Mater Misericordiae University Hospital, Dublin, Ireland. A group of healthly control donors were also recruited. Exclusion criteria consisted of age <18 years and exposure to aspirin or other anti-platelet agents in the preceding 2 weeks.

30 ml of blood was collected via venepuncture with a 21-gauge needle into a 10 mL acid citrate dextrose (ACD) vacutainer and centrifuged (200 xg with no brake for 15 min at room temperature (RT). Platelet rich plasma (PRP) was isolated and the pH adjusted to 6.5 with ACD and supplemented with 1 μM PGE_1_. Platelets were then isolated from plasma by centrifugation (600 × g for 10 min at RT) and platelet-poor-plasma was stored at -80°C.

### Platelet ATP secretion assay

Isolated platelets from severe COVID-19 (n=6), non-severe COVID-19 (n=4) and hospitalised control (n=3) patient groups and healthy controls (n=6) were washed using a modified Tyrode’s buffer (130 mM NaCl, 9 mM NaHCO_3_, 10 mM Tris-HCl, 10 mM Trisodium citrate, 3 mM KCl, 0.81 mM KH_2_PO_4_, 9 mM MgCl_2_ × 6H_2_O; pH 7.4 with ACD) followed by centrifugation (600 xg for 10 min at RT). Platelets were again resuspended in modified Tyrode’s buffer and incubated at 37°C until the ATP secretion assay was performed. Platelet counts were obtained so that the secretion assay results could be normalised to 10^6^ platelets/ml.

ATP secretion was measured using a luminescence-based assay, as previously described (41-44). Agonists and buffer (5μl), were dispensed in duplicate into 96-well plates, together with 35μl of washed platelets and incubated for 3 minutes at 37 °C, with fast orbital shaking. The doses of platelet agonists used to induce platelet ATP secretion were as follows; thrombin: 0.1 U/ml, 0.5 U/ml; U46619: 1 μM, 6.6 μM. Following incubation, 5μl of ATP detecting reagent (chronolume; Labmedics, UK) was dispensed into each well and luminescence was measured immediately using a Perkin Elmer 1420 96-well plate reader. Data was expressed as the amount of ATP secreted, in luminescence arbitrary units (AU), and converted to pmoles ATP released per 10^6^ platelets by comparison with the luminescence recorded from an ATP standard (0.4 mM).

### Enzyme-linked immunosorbent assay (ELISA)

ELISAs for thrombopoietin (DTP00b), P-selectin (DPSE00), and platelet factor 4 (DPF40) were purchased from R&D Systems (Abingdon, UK) and performed according to the manufactures instructions. In brief, undiluted or 1:40 diluted platelet poor plasma (PPP) was added in triplicate to wells for thrombopoietin and PF4, respectively. After incubation, the plates were washed and incubated with the respective conjugate. For P-selectin, 1:20 diluted PPP was added simultaneously with the P-selectin conjugate and incubated. Again, plates were washed and the substrate added. The reaction was stopped with stop solution and the optical density was determined at 450 nm within 30 minutes on a Dynex DS2 (Dynex Technologies, Worthing, UK). Protein quantification was performed using a 4 parametric logistic or log-logit regression.

### Statistical analysis

Statistical analysis was performed using R (version 4.0.0) and SAS Viya© software (version 3.5). Data were tested for normal distribution using a Shapiro-Wilk test. Normally distributed continuous FBC data were presented as mean ± standard deviation and assessed for statistical significance using a one-way ANOVA. Differences in ATP release and protein expression were determined using a two-tailed *t*-test. In both cases, *p*-values were adjusted for multiple comparisons using a Holm-Bonferroni post-hoc test. Categorical data were described as percentages and significance was tested using a Fisher’s exact test. Values below 0.05 were regarded as statistically significant. Data visualisation was performed using SAS Viya©.

## Results

### Mean platelet volume (MPV) and platelet-to-neutrophil ratio (PNR) are associated with disease severity in COVID-19

Clinical laboratory data were available for 54 patients with confirmed SARS-CoV-2 infection. Within this group, 34 patients had developed severe disease requiring transfer to the intensive care unit (ICU) for ventilatory support while the remaining 20 patients did not require critical care support at any stage during the course of their hospital admission. Laboratory parameters relating to these two distinct subgroups of patients with COVID-19 (severe COVID-19, n=34; non-severe COVID-19, n=20) were compared to data from an age and gender-matched cohort of hospitalised medical inpatients without COVID-19 (n=20). Day of admission was defined as the day patients were admitted to the Mater Misericordiae University Hospital.

In agreement with previous studies (45, 46), higher levels of D-dimer were observed on the day of hospital admission among the patients who subsequently developed severe COVID-19 (mean±SD; 4.95±6.36 mg/L) in comparison (*p*=0.0.012) to that observed in the non-severe COVID-19 group (1.01±0.75 mg/L) (Table 1). Furthermore, a significantly higher mean platelet volume (MPV, *p*=0.015) and neutrophil count (*p*=0.013) in addition to a significantly lower platelet-neutrophil ratio (PNR, *p*=0.0067) was noted on the day of admission in the group of patients who subsequently developed a severe disease course relative to that seen in the non-severe group (Fig. 1A). Interestingly these significant differences remained following transfer to intensive care (Fig. 1B).

**Table 1:** Clinical characteristics of hospitalised control patients without COVID-19 on admission and non-severe and severe patients with confirmed COVID-19 on admission. Results are presented as mean±SD.

|  | **Hospitalised**<br>**Controls (n=20)** | **Non-severe**<br>**COVID19**<br>**(n=20)** | **Severe**<br>**COVID19**<br>**(n=34)** | **p-value** |
| --- | --- | --- | --- | --- |
| **Age (years)** | 64.2 ± 17.9 | 69.25 ± 17.7 | 59.4 ± 10.5 | 0.066 |
| **Male, n (%)** | 10 (50%) | 13 (65%) | 21 (62%) | 0.682 |
| **PCT (%)** | 0.262 ± 0.11 | 0.246 ± 0.10 | N/A | 0.626 |
| **P-LCR (%)** | 25.40 ± 9.34 | 23.43 ± 7.57 | N/A | 0.47 |
| **PT (s)** | 13.56 ± 6.92 | 13.26 ± 2.02 | 13.87± 5.47 | 0.886 |
| **aPTT (s)** | 28.84 ± 14.17 | 29.22 ± 1.99 | 33.08± 8.35 | 0.272 |
| **Fibrinogen**<br>**(g/L)** | 5.46 ± 1.72 | 4.65 ± 1.64 | 5.58 ± 1.74 | 0.544 |
| **D-Dimer**<br>**(mg/L)** | 2.53 ± 1.62 | 1.01 ± 0.75 | 4.95 ± 6.36 | **0.027** |

PCT – plateletcrit; PLCR – platelet-large cell ratio; PT – prothrombin time; aPTT – activated partial thromboplastin time

**Figure 1:**
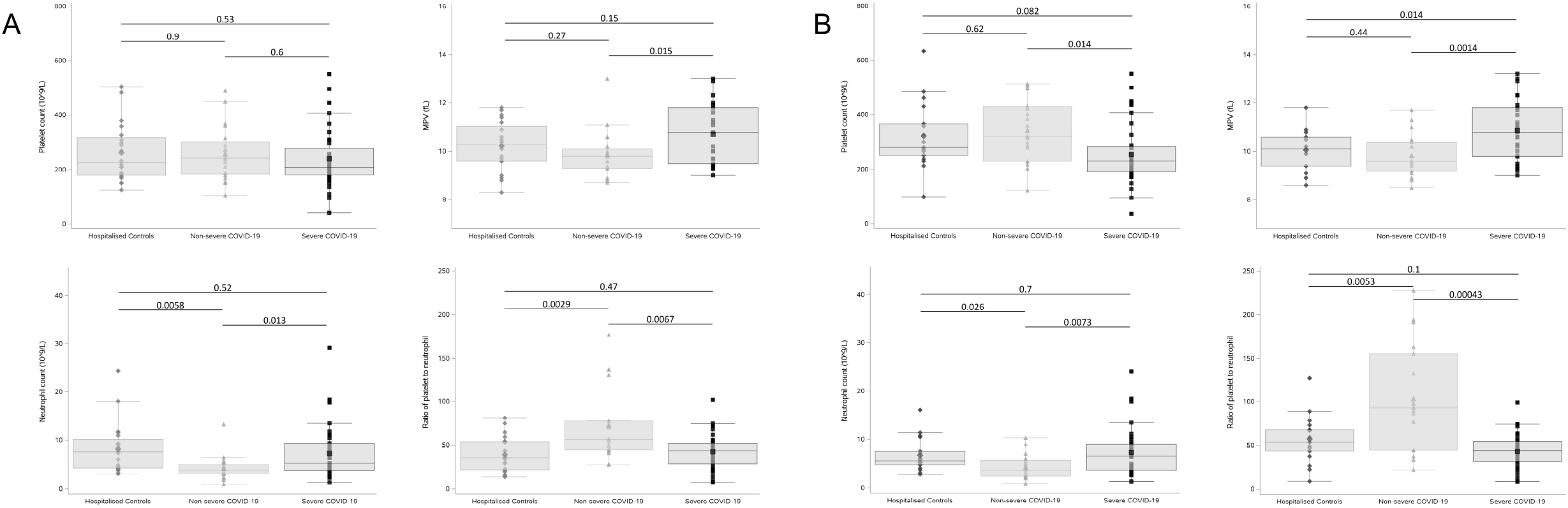
Routinely clinical laboratory parameters are associated with disease severity in COVID-19. Platelet counts, MPV, Neutrophil counts and Platelet-to-neutrophil ratios of patients with severe (n=34) or non-severe (n=20) COVID-19 and hospitalised controls (n=20). **(A)** On the day of admission, patients who subsequently developed severe COVID-19 had significantly higher MPV (*p=*0.015) and neutrophil counts (*p*=0.013) and significantly lower platelet-to-neutrophil ratio (PNR, *p*=0.0067) compared to those who did not subsequently develop severe COVID-19. **(B)** On day 7 of hospitalisation, non-severe COVID-19 patients had a significantly higher platelet count compared to severe COVID-19 patients on the day of transfer to intensive care (*p*=0.014). Statistical analysis was performed using a two-tailed *t*-test and p-values were adjusted for multiple comparisons using a Holm-Bonferroni post hoc test.

While platelet counts did not differ on the day of admission between patients subsequently designated to have had either a severe or non-severe disease course (Fig. 1A), significantly decreased platelet counts were observed among the severe COVID-19 patients at the time of transfer to the ICU relative to the non-severe group (*p*=0.014, Fig. 1B). These findings are in agreement with previous studies, which have suggested that a lower platelet count is associated with severe COVID-19 (3, 47-49). An increased neutrophil-to-lymphocyte ratio (NLR) in severe cases relative to non-severe was also detected at the time of hospital admission (another previously reported predictor of disease severity) (Supplementary Fig. 1) (47, 50, 51).

### Agonist-induced ADP release is dramatically higher in COVID-19 patients compared with non-COVID-19 hospitalized patients

Recently published data have suggested a crucial mechanistic role for platelets in the pathophysiology of COVID-19 (18, 32, 52). We assessed dense granule release following platelet activation with various platelet agonists. ATP secretion was used as a surrogate marker for dense granule secretion, given that the ratio of ADP to ATP in platelet dense granules is a constant 2:3 ratio (53).

Platelets from individuals within each group were isolated and activated with low and high concentrations of thrombin and U46619. Platelets from COVID-19 patients exhibited significantly higher levels of dense granule secretion when compared to both control cohorts (HeC and HoC) with no significant difference between severe and non-severe COVID-19 patients (Fig. 2). Low doses of thrombin (0.1 U/ml) and U46619 (1 μM) induced minimal activation in platelets isolated from healthy and hospitalised control patients. In contrast, dramatically increased dense granule secretion was observed upon platelet activation in patients with COVID-19 (Fig. 2A, 2C and Supplementary Table 2). Platelet stimulation with 0.1U/mL thrombin increased ADP-release 3-fold in COVID-19 patients compared with HeC and HoC (Fig. 2A and Supplementary Table 2). Crucially, platelet activation with the thromboxane A_2_ receptor agonist U46619 (1µM) induced a 28-fold increase in ADP-release in COVID-19 positive patients compared to HeC and strikingly, an 89-fold increase between COVID-19 positive patients and HoC (Fig. 2C and Supplementary Table 2). Platelet stimulation with high-dose thrombin (0.5 U/ml) and U46619 (6.6 μM) induced a 2-fold and 10-fold increase respectively, in dense granule secretion from COVID-19 patients compared to controls (Fig. 2B, 2D and Supplementary Table 2). These data suggest that platelets from COVID-19 patients are more responsive to low dose agonist stimulation when compared to control platelets, potentially indicating a reduced activation threshold in COVID-19 patients.

**Figure 2:**
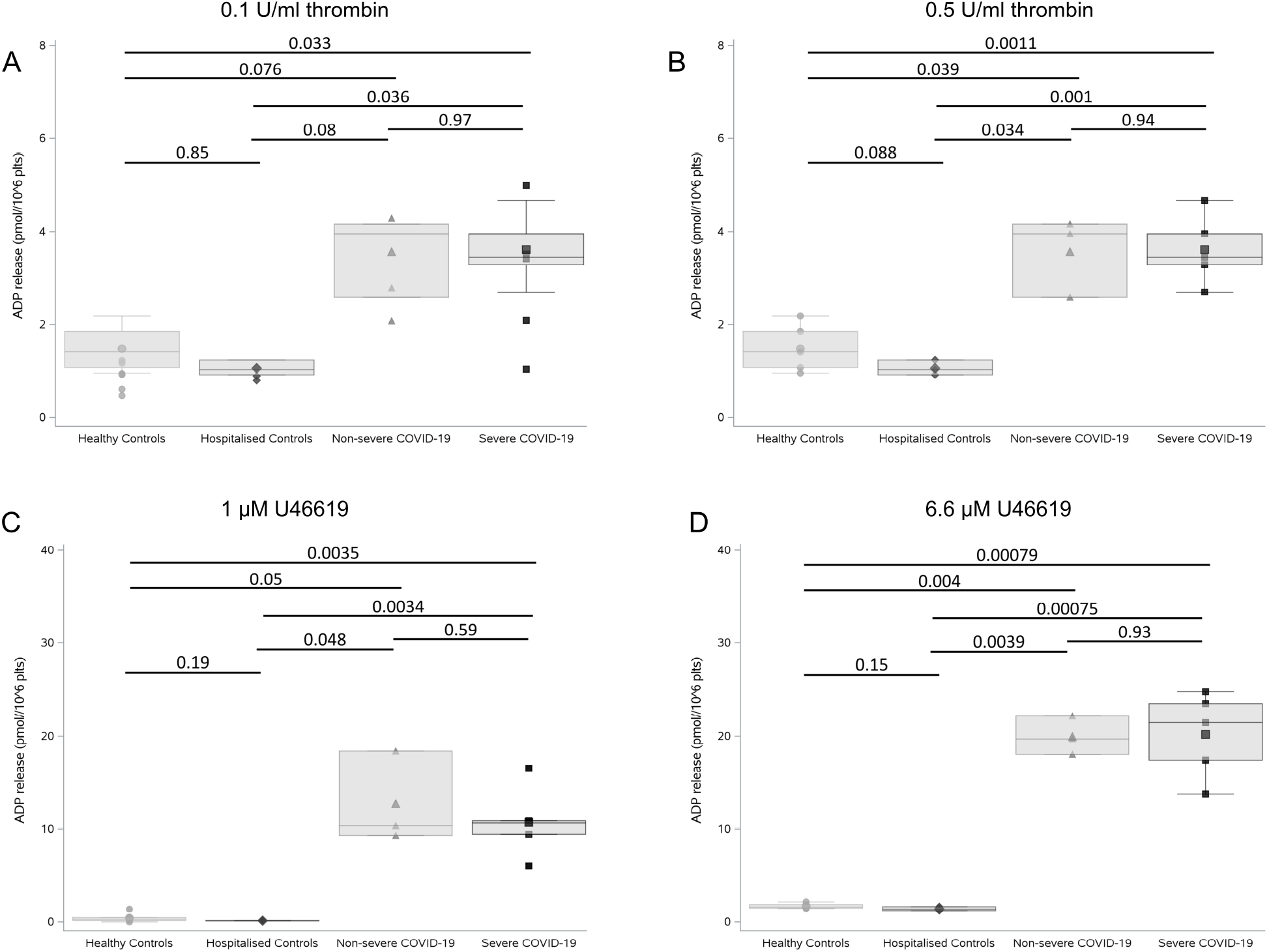
Agonist-induced ADP release was dramatically higher in COVID-19 patients compared with non-COVID-19 hospitalized patients. Platelet dense granule release was measured in duplicate in severe (n=5) and non-severe (n=4) COVID-19 patients compared to hospitalised (n=3) and healthy controls (n=6). Platelets were stimulated with 0.1 U/ml thrombin **(A)**, 0.5 U/ml thrombin **(B)**, 1 µM U46619 **(C)**, and 6.6 µM U46619 **(D)** and ATP release (surrogate for ADP) was measured using a Chronolume luciferase assay. ADP release is expressed as pmol/10^6^ platelets. Statistical analysis was performed using a two-tailed *t*-test and p-values were adjusted for multiple comparisons using a Holm-Bonferroni post hoc test.

### Circulating levels of PF4, sP-selectin and TPO are significantly elevated in COVID-19

To further characterise this hyperactive platelet phenotype, circulating plasma markers of platelet activity were measured in each of the above study groups (n=6 per group; platelet-poor plasma was available from an additional 2 non-severe COVID patients and an additional 3 hospitalised controls). Circulating levels of platelet factor 4 (PF4) were substantially elevated in COVID-19 patients when compared to both hospitalised (HoC) and healthy controls (HeC) (*p*=0.012 and *p*=0.0033, respectively; Fig. 3A). Nonetheless circulating PF4 levels could not differentiate between severe and non-severe COVID-19 patient cohorts (*p*=0.85). On the other hand, circulating levels of soluble P-selectin (sCD62P) also increased in COVID-19 patients when compared to our control cohorts (Fig. 3B) and intriguingly, could differentiate between our non-severe and severe COVID-19 cohorts (*p*=0.037), suggesting that elevated sP-selectin may be associated with disease severity.

**Figure 3:**
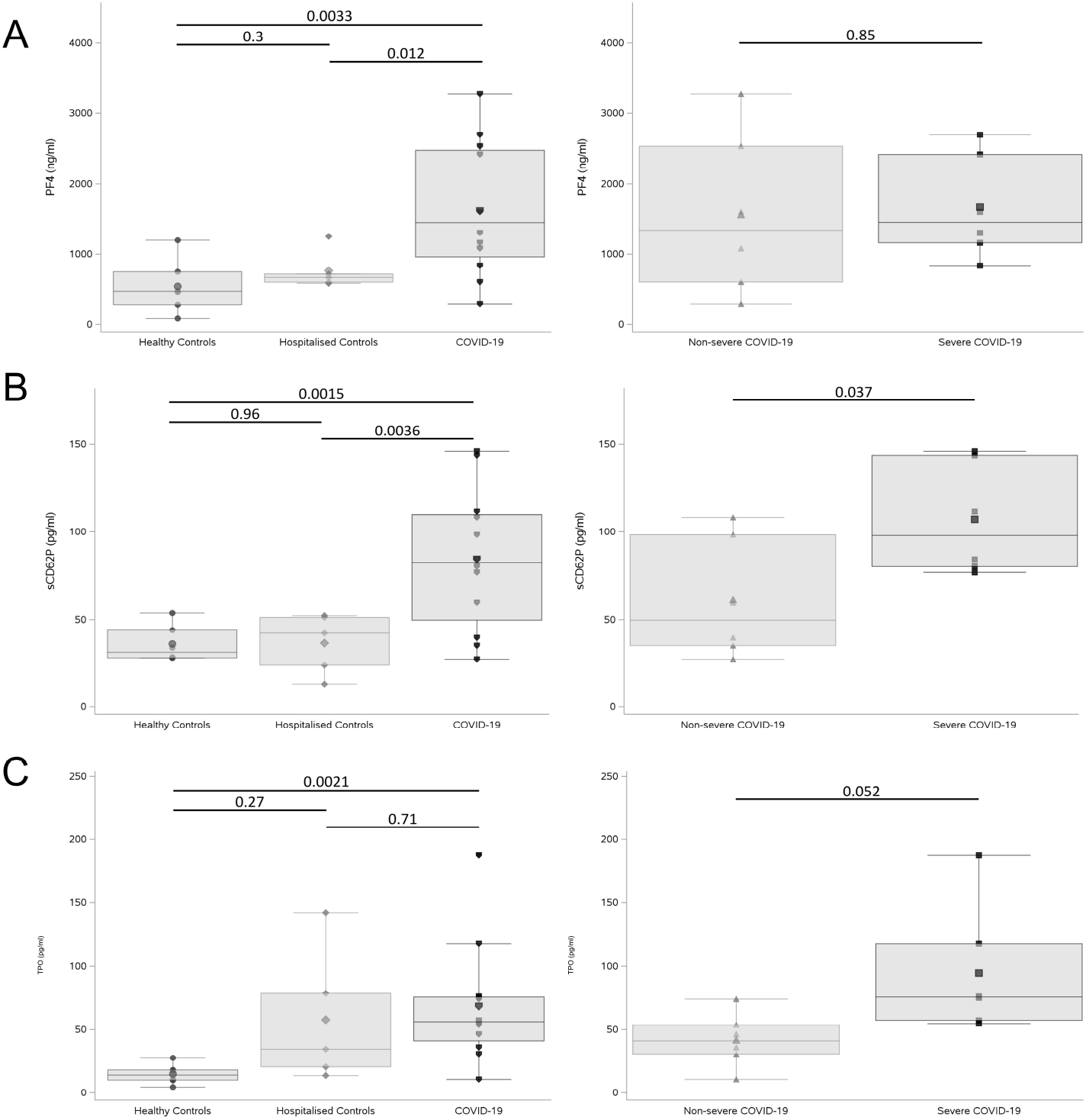
Circulating levels of PF4, sP-selectin levels and TPO were significantly elevated in COVID-19. **(A)** Platelet factor 4 (PF), **(B)** soluble P-selectin (sCD62P), and **(C)** thrombopoietin (TPO) plasma levels from patients with severe (n=6) and non-severe (n=6) COVID-19 compared to hospitalized (n=6) and healthy (n=6) controls were measured in triplicate by ELISA. Statistical analysis was performed using a two-tailed *t*-test and p-values were adjusted for multiple comparisons using a Holm-Bonferroni post hoc test.

Increased thrombopoetin (TPO) levels have been reported in SARs-CoV infection and more recently in COVID-19 (only in comparison to healthy controls) (54). Within our study population, TPO levels were found to be significantly higher among patients with COVID-19 (combined severe and non-severe) compared to HeC (*p*=0.0021; Fig. 3C). No significant difference in circulating TPO levels was noted between COVID-19 and HoC patients (*p*=0.71; Fig. 3C). Interestingly, TPO levels between our severe and non-severe COVID-19 cohorts showed a clear trend towards elevated circulating TPO in severe COVID-19 infection (*p*=0.052; Fig. 3C).

## Discussion

We have demonstrated that distinct platelet-related patterns in full blood count and other clinical laboratory parameters appear to be associated with the development of a more severe disease course in our cohort of COVID-19 patients, reflecting other recently published data (2, 47, 55). Therewithal, we also report novel findings which indicate that COVID-19 is associated with abnormal platelet reactivity. Our observations are likely to be of significant clinical and translational relevance in view of the uncertainty that currently exists with regards to the aetiology of thrombotic risk in COVID-19 and the optimal strategy for prevention and treatment of thrombosis in this population.

In addition to the previously reported observations that neutrophilia, lymphopenia, elevated D-dimer, elevated neutrophil-to-lymphocyte ratio and elevated platelet-to-lymphocyte ratio appear to be associated with severe disease, we have reported that within our cohort a significantly lower platelet-to-neutrophil ratio was observed on the day of admission among the COVID-19 patients who subsequently progressed to require critical care support (5, 11, 12, 47, 56). While platelet counts did not specifically differ between groups on day of hospitalisation, we did observe increased levels of thrombopoietin in all COVID-19 positive patients, potentially indicating the presence of a stimulus for increased thrombopoeisis in this group. Moreover, a greater MPV was observed on the day of hospital admission in the group of COVID-19 patients who subsequently developed a severe disease course requiring transfer to intensive care. Enlarged platelets express more platelet activation markers (57) and contain more granules (58) and the release of such granular contents (e.g. thromboxane, ADP and β-thromboglobulin) (59) can drive further platelet activation.

Other investigators have demonstrated increased platelet aggregation in response to low-dose agonist stimulation in COVID-19 patients as well as increased platelet thromboxane generation (32, 60). In line with these findings, we observed increased platelet responsiveness (ADP release) to low-dose agonist stimulation, within our population of COVID-19 patients. Intriguingly, previous reports have demonstrated similar patterns of enhanced platelet reactivity in response to subthreshold concentrations of agonists in the presence of dengue virus nonstructural protein 1 (34, 61), potentially indicating a virus-induced sensitisation of platelets. As ADP/ATP and thromboxane A2 are locally-acting autocrine agonists which rapidly amplify platelet activation triggering more release of platelet granule contents, our findings provide further evidence that platelets from COVID-19 patients are more reactive, a phenomenon which may contribute to thrombotic risk (4-10).

The circulating levels of the platelet activation markers PF4 and sP-selectin were highly elevated among our cohort of COVID-19 positive patients compared to controls which futher supports the hypothesis that COVID-19 is associated with enhanced platelet activation. The elevated levels of TPO in COVID-19 also aligned with previous findings (32). While circulating levels of PF4 did not differentiate between severe and non-severe COVID-19 cases, sP-selectin was higher among the severe COVID-19 group, in keeping with previous reports (60, 62). Other investigators have also reported that sP-selectin levels were elevated in COVID-19 complicated by acute respiratory distress syndrome (ARDS) (63), with even higher levels detected in patients who subsequently died (64). The mechanism underlying platelet activation in COVID-19 has been speculated to involve MAP kinase pathway (32) and/or PKCd (60) activation, however the initial trigger remains unidentified. Crucially, Zaid *et al*. detected mRNA for angiotensin-converting enzyme 2 (ACE2), the membrane receptor that facilitates SARS-CoV-2 internalisation, in platelets (60), but it remains to be determined if platelets are directly infected by the virus. Potential alternative pathways may include internalisation of the virus or the virus genome or binding to other cell surface receptors (e.g. FcγRIIA or Dendritic Cell-Specific Intercellular adhesion molecule-3-Grabbing Non-integrin), as observed with other viral infections (33, 65-68).

This study has a number of limitations. Firstly, our cohort of patients is relatively small, a factor which (at least in part) is likely to be related to the success of the Irish healthcare system in suppressing transmission of the virus during the early stages of the pandemic. Additionally, given the public health restrictions that were in place during the period of this study, recruitment of healthy controls was substantially impeded and consequently the available healthy controls were not appropriately matched for age relative to our COVID-19 group and hospitalised control group. Due to the small sample size, adjustment for confounding factors was not possible.

In conclusion, it has been apparent since the early stages of the COVID-19 pandemic that derangements of haemostasis represent a hallmark of this disease and appear to be associated with significant morbidity (12, 69). Several investigators have reported data suggesting that rates of venous thromboembolism are high in this population and that these thrombotic complications appear to occur despite compliance with standard VTE prevention protocols (5, 7). Reports of unusual and devastating cases of arterial thrombosis are also emerging (18, 22-24). The safety and efficacy of conventional anti-platelet and anti-coagulant treatment strategies in the management of this thrombotic risk is becoming a major focus of translational and clinical interest. The aetiology of hypercoagulability in COVID-19 is likely multi-factorial but it is presumed to be driven by the marked inflammatory response which arises following infection (70, 71). It remains to be determined whether standard thrombosis prevention strategies require modification in order to be effective in the prevention of ‘immunothrombosis’ in COVID-19 or if prevention strategies should focus instead on addressing the ‘upstream’ inflammatory response driving coagulation activation. Our data indicate that platelets from patients infected with COVID-19 display a hyperactive phenotype, a factor which may contribute to thrombotic risk. Consequently, further characterisation of platelet dysfunction in COVID-19 and the evaluation of anti-platelet therapy as an adjunct to current thrombosis prevention measures is clearly warranted.

## Data Availability

All data, analytic methods and study materials supporting the findings of this study are provided in the manuscript, supplementary material and available from the corresponding authors upon reasonable request.

## Acknowledgements

We thank John Crumlish, Áine Lennon, Jane Culligan, Charlotte Prior, Wendy Keogh and Julie McAndrew of the Department of Pathology, Mater Misericordiae University Hospital, for the help in preparing blood samples and ELISA experiments. We also thank Annette Wallace of Conway Stores for her invaluable help. We also thank all patients who gave consent and donated blood to this study. Finally, we thank all frontline healthcare workers in Ireland for their dedication and professionalism during the COVID-19 pandemic.

## Author Contribution

S.P. Comer, P.B. Szklanna, L. Weiss, F. Ní Áinle, B. Kevane and P.B. Maguire designed the research and wrote the paper. S.P. Comer, P.B. Szklanna and L. Weiss performed experiments, and analysed data. S. Cullen performed the ATP secretion assay. S. Cullivan, S. Kelliher, F Ní Áinle and B. Kevane identified and recruited patients to the study. A. Smolenski, N. Moran, C. Murphy, K. O’Reilly, A. G. Cotter, B. Marsh, S. Gaine, P. Mallon and B. McCullagh contributed to research design and paper writing. H. Altaie and J. Curran contributed data analytics support and expertise.

## Appendix

### Additional investigators of the COCOON Study (all in Ireland unless stated)

Peter Doran (Scientific Director, UCD Clinical Research Centre), Jack Lambert (Dept. of Infectious Diseases, Mater Misericordiae University Hospital/UCD School of Medicine), Gerard Sheehan (Dept. of Infectious Diseases, Mater Misericordiae University Hospital/UCD School of Medicine), Tara McGinty (Dept.of Infectious Diseases, Mater Misericordiae University Hospital/UCD School of Medicine), Aoife Cotter (Dept. of Infectious Diseases, Mater Misericordiae University Hospital/UCD School of Medicine), Colman O’Loughlin (Dept.of Critical Care, Mater Misericordiae University Hospital), Jennifer Hastings (Dept. of Critical Care, Mater Misericordiae University Hospital), Mairead Hayes (Dept. of Critical Care, Mater Misericordiae University Hospital), Margaret Doherty (Dept. of Critical Care, Mater Misericordiae University Hospital), Frances Colreavy (Dept. of Critical Care, Mater Misericordiae University Hospital), Ian Conraic-Martin (Dept. of Critical Care, Mater Misericordiae University Hospital), Peter McMahon (Dept. of Radiology, Mater Misericordiae University Hospital/UCD School of Medicine) Leo Lawler (Dept. of Radiology, Mater Misericordiae University Hospital/UCD School of Medicine), Dermot O’Callaghan (Dept. of Respiratory Medicine, Mater Misericordiae University Hospital), Tomas Breslin (Dept. of Emergency Medicine, Mater Misericordiae University Hospital/UCD School of Medicine), Cian McDermott (Dept. of Emergency Medicine; Mater Misericordiae University Hospital/UCD School of Medicine), Robert Turner (Dept. of Critical Care Medicine; Mater Misericordiae University Hospital/UCD School of Medicine), John Crumlish (Laboratory Manager, Mater Misericordiae University Hospital), Aine Lennon (Dept. of Haematology, Mater Misericordiae University Hospital), Wendy Keogh (Dept. of Haematology, Mater Misericordiae University Hospital), Jane Culligan (Dept. of Haematology, Mater Misericordiae University Hospital), Siobhán McClean (UCD School of Biomolecular & Biomedical Science/UCD Conway Institute), Patrick Mallon (Dept. of Infectious Diseases, St Vincent’s University Hospital/UCD School of Medicine), Jennifer Donnelly (Dept. of Obstetrics, Rotunda Hospital), Mary Higgins (Dept. of Obstetrics, National Maternity Hospital), Annemarie O’Neill (Founder, Thrombosis Ireland Patient Organization), Rachel Rosovsky (Massachusetts General Hospital/Harvard Medical School, Massachusetts, USA), Karen Schreiber (Dept. of Rheumatology, Copenhagen University Hospital, Denmark), Cliona Ní Cheallaigh (Dept. of Infectious Diseases/Director of Inclusion Medicine, St James’s Hospital/TCD School of Medicine), Mary Cushman (Dept. of Medicine & Pathology, Vermont Medical Center & Larner College of Medicine, University of Vermont, Vermont, USA), Saskia Middeldorp (Academic Medical Centre, Amsterdam, the Netherlands), Peter Juni (Dept. of Medicine, St Michaels Hospital & University of Toronto, Canada), Michelle Sholzberg (Dept. of Medicine, St Michaels Hospital & University of Toronto, Canada).

